# Predicting the disease outcome in COVID-19 positive patients through Machine Learning: a retrospective cohort study with Brazilian data

**DOI:** 10.1101/2020.06.26.20140764

**Authors:** Fernanda Sumika Hojo de Souza, Natália Satchiko Hojo-Souza, Edimilson Batista dos Santos, Cristiano Maciel da Silva, Daniel Ludovico Guidoni

## Abstract

The first officially registered case of COVID-19 in Brazil was on February 26, 2020. Since then, the situation has worsened with more than 672, 000 confirmed cases and at least 36, 000 reported deaths at the time of this writing. Accurate diagnosis of patients with COVID-19 is extremely important to offer adequate treatment, and avoid overloading the healthcare system. Characteristics of patients such as age, comorbidities and varied clinical symptoms can help in classifying the level of infection severity, predict the disease outcome and the need for hospitalization. Here, we present a study to predict a poor prognosis in positive COVID-19 patients and possible outcomes using machine learning. The study dataset comprises information of 13, 690 patients concerning closed cases due to cure or death. Our experimental results show the disease outcome can be predicted with a ROC AUC of 0.92, Sensitivity of 0.88 and Specificity of 0.82 for the best prediction model. This is a preliminary retrospective study which can be improved with the inclusion of further data. Conclusion: Machine learning techniques fed with demographic and clinical data along with comorbidities of the patients can assist in the prognostic prediction and physician decision-making, allowing a faster response and contributing to the non-overload of healthcare systems.

## 1 Introduction

A new coronavirus with a high efficiency in infecting humans emerged in the Wuhan city (Hubei Province, China) in December 2019. The disease, named COVID-19, is caused by the severe acute respiratory syndrome coronavirus 2 (SARS-CoV-2) being highly contagious [1, 2]. The virus spread quickly outside China and the World Health Organization (WHO) recognized the outbreak as a pandemic in March 2020 [3]. To date, nearly 7 million cases have been confirmed and more than 400, 000 deaths have been reported from SARS-Co-2 infection, reaching 216 countries [4].

Despite high transmissibility, the disease spectrum is diverse, ranging from asymptomatic cases to extremely severe conditions. SARS-CoV-2 infection is characterized by fever, generalized weakness, dry cough, headache, dyspnoea, myalgia, as well as leukopenia, lymphocytopenia, neutrophilia, elevated levels of C-reactive protein, D-dimer, and inflammatory cytokines [5, 6, 7] and loss of smell and taste in the early stage of infection [8]. However, the status can quickly evolve to acute respiratory distress syndrome (ARDS), cytokine storm, coagulation dysfunction, acute cardiac injury, acute kidney injury, and multi-organ dysfunction if the disease is not resolved, resulting in patient death [9, 10].

Elderly people and those with comorbidities such as diabetes and cardiovascular disease are more likely to progress to severe conditions [7]. Obesity has also been linked to an increased likelihood of severe COVID-19 [11]. In this context, a three-stage classification system was proposed according to the degree of severity of the disease [12]. The initial stage is characterized by nonspecific clinical symptoms such as malaise, cough and fever. The diagnosis can be made in a respiratory sample to detect the presence of SARS-CoV-2 by RT-PCR and blood tests can reveal lymphopenia and neutrophilia. The second stage is characterized by viral lung disease and localized inflammation usually associated with hypoxia, requiring hospitalization and mechanical ventilation. However, there is a number of cases that progress to a more severe third stage of the disease, characterized by high levels of inflammatory biomarkers and severe systemic inflammation. In this phase, the occurrence of shock and generalized collapse of the organs is large and the prognosis for recovery is poor.

So far, there is no vaccine or even effective therapeutic drugs for the treatment of COVID-19 [13]. Therefore, quarantine and social distancing have been recommended as a measure to reduce the rate of infection aiming not to exceed the capacity of health systems to provide care.

Currently, COVID-19 is on the rise in Latin American countries [14], whose health systems may not support the care of all seriously infected people. Lack of beds, ventilators in Intensive Care Units (ICUs) and Personal Protective Equipment (PPE) by health care workers restrain the treatment of severe cases. Faced with these challenges, identifying those patients with hospitalization priority is a crucial aspect in order to optimize care and promote a reduction of deaths.

At present, Brazil is experiencing a critical situation with more than 672, 000 confirmed cases and at least 36, 000 deaths (WHO, 2020). Brazil has become the epicenter of the pandemic, which expands to interior cities, most of which do not have ICUs.

The rapid spread of COVID-19 associated with the lack of a vaccine and effective therapeutic measures has accelerated the use of artificial intelligence and machine learning on different fronts such as viral dissemination pattern, rapid and accurate diagnosis, development of new therapeutic approaches and identification of people most susceptible to the disease [15].

The aim of the present study is to make a prognosis or early identification of patients at increased risk of developing severe COVID-19 symptoms using an available database from the Espírito Santo Brazilian State. Espírito Santo has an estimated population of 4.06 million inhabitants [16] and on May 30th, 2020 had registered 13, 690 confirmed cases of COVID-19. Using machine learning techniques, a classification problem can be solved aiming to predict the disease outcome for positive COVID-19 patients, based on individual information, in addition to comorbidities and varied clinical symptoms. We show that it is possible to predict the outcome of the individual’s condition with a ROC AUC of 0.92, Sensitivity of 0.88 and Specificity of 0.82. This process can be of great importance in helping decision-making within hospitals, since resources are becoming limited every day. Patients classified as having a more severe condition can be prioritized in this case.

## 2 Data and Methods

This is a retrospective cohort study that did not directly involve patients and does not require approval by an ethics committee. The database used is publicly available on the Espírito Santo state portal [17]. Two sets of data are used in our study, say the training cohort and the validation cohort. The database was downloaded twice, on May 23rd, 2020 and May 30th, 2020. Information regarding 13, 690 patients who tested positive for COVID-19 comprises the last database, along with the outcome of each case. As the main objective of the present work is to predict the disease outcome of patients infected by the virus, only closed cases (due to death or cure) are used, comprising 4, 826 and 3, 617 patients in the training cohort and validation cohort, respectively. Additional information on cleaning and preparing the data is provided below, followed by the machine learning methods employed.

### 2.1 Data Cleaning and Preparation

The dataset includes individual basic information such as gender and age range, symptoms, comorbidities and a recent travelling history. A notification status of each entry in the database is said to be *closed* or *open*, since the data is updated daily as new information becomes available. Thus, only data whose status is *closed* were considered, as they are those that have the outcome of the case: *cure* or *death*. Cases whose outcome is unknown have been disregarded.

The city of origin of the patients and the neighborhood of residence are also available in the database. We considered that this information would not be very relevant to the problem under study, and we decided not to include such data in our datasets due to its high variability in values and possible noise generation in the experiments.

Therefore, based on the data available at the source, we end up our datasets with the following information: confirmation criteria, age range, gender, race/color, education, fever, respiratory distress, cough, runny nose, sore throat, diarrhea, headache, pulmonary comorbidity, cardiac comorbidity, kidney comorbidity, diabetes comorbidity, smoking comorbidity, obesity comorbidity, hospitalization, travel in Brazil and international travel. All of these variables comprise categorical variables, taking a value from a finite discrete set. Tables 1 and 2 details the dataset variables for all training and validation patients, respectively, showing their distribution among the categories, as well as separated by the outcome, i.e., cure or death.

**Table 1:**
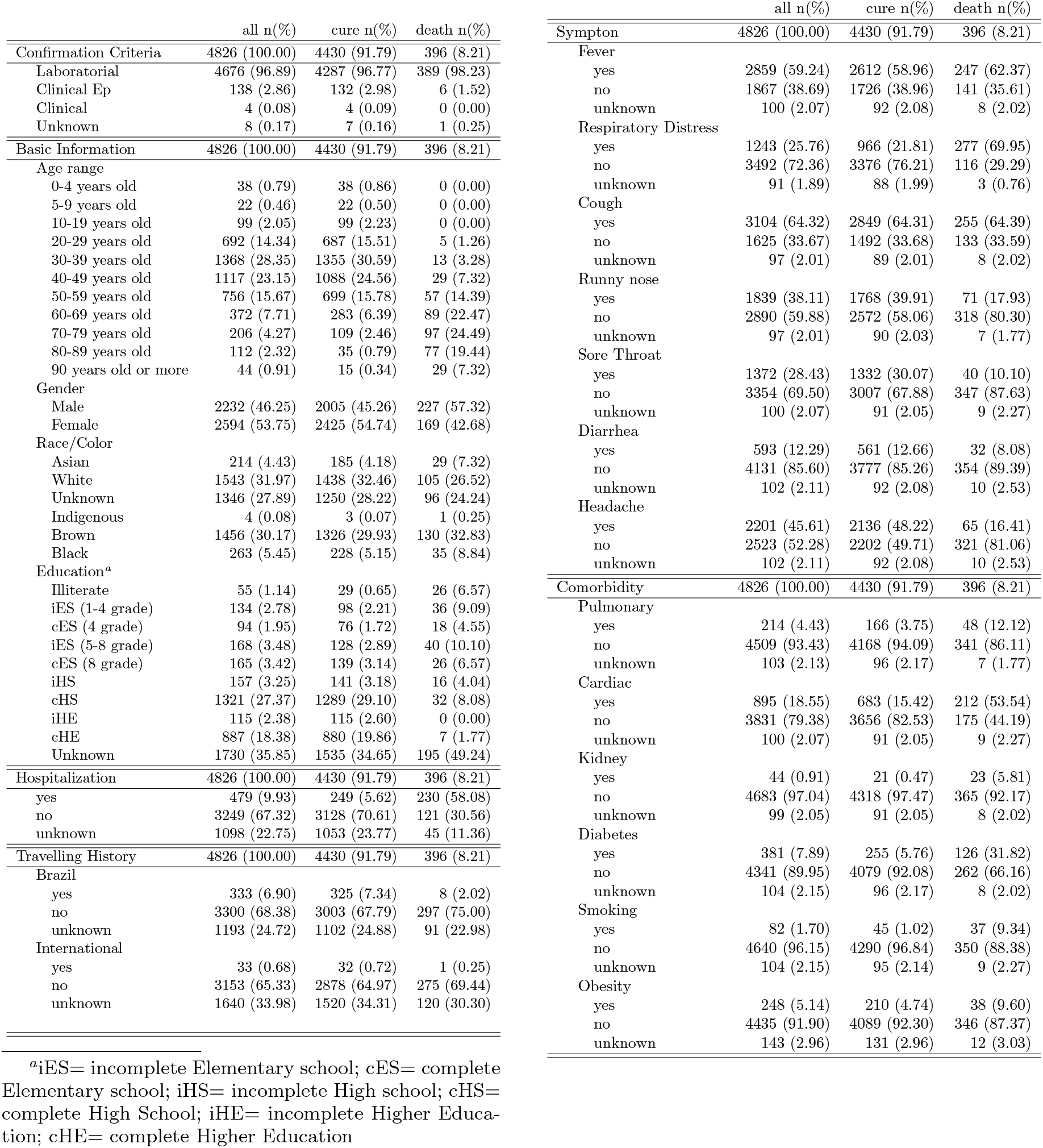
Demographic data and clinical characteristics of the study population Training Dataset (Feb 29th - May 23rd)

|  | all n(%) | cure n(%) | death n(%) |
| --- | --- | --- | --- |
| Confirmation Criteria | 4826 (100.00) | 4430 (91.79) | 396 (8.21) |
| Laboratorial | 4676 (96.89) | 4287 (96.77) | 389 (98.23) |
| Clinical Ep | 138 (2.86) | 132 (2.98) | 6 (1.52) |
| Clinical | 4 (0.08) | 4 (0.09) | 0 (0.00) |
| Unknown | 8 (0.17) | 7 (0.16) | 1 (0.25) |
| Basic Information | 4826 (100.00) | 4430 (91.79) | 396 (8.21) |
| Age range |  |  |  |
| 0-4 years old | 38 (0.79) | 38 (0.86) | 0 (0.00) |
| 5-9 years old | 22 (0.46) | 22 (0.50) | 0 (0.00) |
| 10-19 years old | 99 (2.05) | 99 (2.23) | 0 (0.00) |
| 20-29 years old | 692 (14.34) | 687 (15.51) | 5 (1.26) |
| 30-39 years old | 1368 (28.35) | 1355 (30.59) | 13 (3.28) |
| 40-49 years old | 1117 (23.15) | 1088 (24.56) | 29 (7.32) |
| 50-59 years old | 756 (15.67) | 699 (15.78) | 57 (14.39) |
| 60-69 years old | 372 (7.71) | 283 (6.39) | 89 (22.47) |
| 70-79 years old | 206 (4.27) | 109 (2.46) | 97 (24.49) |
| 80-89 years old | 112 (2.32) | 35 (0.79) | 77 (19.44) |
| 90 years old or more | 44 (0.91) | 15 (0.34) | 29 (7.32) |
| Gender |  |  |  |
| Male | 2232 (46.25) | 2005 (45.26) | 227 (57.32) |
| Female | 2594 (53.75) | 2425 (54.74) | 169 (42.68) |
| Race/Color |  |  |  |
| Asian | 214 (4.43) | 185 (4.18) | 29 (7.32) |
| White | 1543 (31.97) | 1438 (32.46) | 105 (26.52) |
| Unknown | 1346 (27.89) | 1250 (28.22) | 96 (24.24) |
| Indigenous | 4 (0.08) | 3 (0.07) | 1 (0.25) |
| Brown | 1456 (30.17) | 1326 (29.93) | 130 (32.83) |
| Black | 263 (5.45) | 228 (5.15) | 35 (8.84) |
| Education<sup>a</sup> |  |  |  |
| Illiterate | 55 (1.14) | 29 (0.65) | 26 (6.57) |
| iES (1-4 grade) | 134 (2.78) | 98 (2.21) | 36 (9.09) |
| cES (4 grade) | 94 (1.95) | 76 (1.72) | 18 (4.55) |
| iES (5-8 grade) | 168 (3.48) | 128 (2.89) | 40 (10.10) |
| cES (8 grade) | 165 (3.42) | 139 (3.14) | 26 (6.57) |
| iHS | 157 (3.25) | 141 (3.18) | 16 (4.04) |
| cHS | 1321 (27.37) | 1289 (29.10) | 32 (8.08) |
| iHE | 115 (2.38) | 115 (2.60) | 0 (0.00) |
| cHE | 887 (18.38) | 880 (19.86) | 7 (1.77) |
| Unknown | 1730 (35.85) | 1535 (34.65) | 195 (49.24) |
| Hospitalization | 4826 (100.00) | 4430 (91.79) | 396 (8.21) |
| yes | 479 (9.93) | 249 (5.62) | 230 (58.08) |
| no | 3249 (67.32) | 3128 (70.61) | 121 (30.56) |
| unknown | 1098 (22.75) | 1053 (23.77) | 45 (11.36) |
| Travelling History | 4826 (100.00) | 4430 (91.79) | 396 (8.21) |
| Brazil |  |  |  |
| yes | 333 (6.90) | 325 (7.34) | 8 (2.02) |
| no | 3300 (68.38) | 3003 (67.79) | 297 (75.00) |
| unknown | 1193 (24.72) | 1102 (24.88) | 91 (22.98) |
| International |  |  |  |
| yes | 33 (0.68) | 32 (0.72) | 1 (0.25) |
| no | 3153 (65.33) | 2878 (64.97) | 275 (69.44) |
| unknown | 1640 (33.98) | 1520 (34.31) | 120 (30.30) |
| Symptom | 4826 (100.00) | 4430 (91.79) | 396 (8.21) |
| Fever |  |  |  |
| yes | 2859 (59.24) | 2612 (58.96) | 247 (62.37) |
| no | 1867 (38.69) | 1726 (38.96) | 141 (35.61) |
| unknown | 100 (2.07) | 92 (2.08) | 8 (2.02) |
| Respiratory Distress |  |  |  |
| yes | 1243 (25.76) | 966 (21.81) | 277 (69.95) |
| no | 3492 (72.36) | 3376 (76.21) | 116 (29.29) |
| unknown | 91 (1.89) | 88 (1.99) | 3 (0.76) |
| Cough |  |  |  |
| yes | 3104 (64.32) | 2849 (64.31) | 255 (64.39) |
| no | 1625 (33.67) | 1492 (33.68) | 133 (33.59) |
| unknown | 97 (2.01) | 89 (2.01) | 8 (2.02) |
| Runny nose |  |  |  |
| yes | 1839 (38.11) | 1768 (39.91) | 71 (17.93) |
| no | 2890 (59.88) | 2572 (58.06) | 318 (80.30) |
| unknown | 97 (2.01) | 90 (2.03) | 7 (1.77) |
| Sore Throat |  |  |  |
| yes | 1372 (28.43) | 1332 (30.07) | 40 (10.10) |
| no | 3354 (69.50) | 3007 (67.88) | 347 (87.63) |
| unknown | 100 (2.07) | 91 (2.05) | 9 (2.27) |
| Diarrhea |  |  |  |
| yes | 593 (12.29) | 561 (12.66) | 32 (8.08) |
| no | 4131 (85.60) | 3777 (85.26) | 354 (89.39) |
| unknown | 102 (2.11) | 92 (2.08) | 10 (2.53) |
| Headache |  |  |  |
| yes | 2201 (45.61) | 2136 (48.22) | 65 (16.41) |
| no | 2523 (52.28) | 2202 (49.71) | 321 (81.06) |
| unknown | 102 (2.11) | 92 (2.08) | 10 (2.53) |
| Comorbidity | 4826 (100.00) | 4430 (91.79) | 396 (8.21) |
| Pulmonary |  |  |  |
| yes | 214 (4.43) | 166 (3.75) | 48 (12.12) |
| no | 4509 (93.43) | 4168 (94.09) | 341 (86.11) |
| unknown | 103 (2.13) | 96 (2.17) | 7 (1.77) |
| Cardiac |  |  |  |
| yes | 895 (18.55) | 683 (15.42) | 212 (53.54) |
| no | 3831 (79.38) | 3656 (82.53) | 175 (44.19) |
| unknown | 100 (2.07) | 91 (2.05) | 9 (2.27) |
| Kidney |  |  |  |
| yes | 44 (0.91) | 21 (0.47) | 23 (5.81) |
| no | 4683 (97.04) | 4318 (97.47) | 365 (92.17) |
| unknown | 99 (2.05) | 91 (2.05) | 8 (2.02) |
| Diabetes |  |  |  |
| yes | 381 (7.89) | 255 (5.76) | 126 (31.82) |
| no | 4341 (89.95) | 4079 (92.08) | 262 (66.16) |
| unknown | 104 (2.15) | 96 (2.17) | 8 (2.02) |
| Smoking |  |  |  |
| yes | 82 (1.70) | 45 (1.02) | 37 (9.34) |
| no | 4640 (96.15) | 4290 (96.84) | 350 (88.38) |
| unknown | 104 (2.15) | 95 (2.14) | 9 (2.27) |
| Obesity |  |  |  |
| yes | 248 (5.14) | 210 (4.74) | 38 (9.60) |
| no | 4435 (91.90) | 4089 (92.30) | 346 (87.37) |
| unknown | 143 (2.96) | 131 (2.96) | 12 (3.03) |

<sup>a</sup>iES= incomplete Elementary school; cES= complete Elementary school; iHS= incomplete High school; cHS= complete High School; iHE= incomplete Higher Education; cHE= complete Higher Education

**Table 2:** Demographic data and clinical characteristics of the study population Validation Dataset (May 24th - May 30th)

|  | all n(%) | cure n(%) | death n(%) |
| --- | --- | --- | --- |
| Confirmation Criteria | 3617 (100.00) | 3396 (93.89) | 221 (6.11) |
| Laboratorial | 3479 (96.18) | 3259 (95.97) | 220 (99.55) |
| Clinical Ep | 108 (2.99) | 108 (3.18) | 0 (0.00) |
| Clinical | 12 (0.33) | 11 (0.32) | 1 (0.45) |
| Unknown | 18 (0.50) | 18 (0.53) | 0 (0.00) |
| Basic Information | 3617 (100.00) | 3396 (93.89) | 221 (6.11) |
| Age range |  |  |  |
| 0-4 years old | 30 (0.83) | 29 (0.85) | 1 (0.45) |
| 5-9 years old | 23 (0.64) | 23 (0.68) | 0 (0.00) |
| 10-19 years old | 67 (1.85) | 67 (1.97) | 0 (0.00) |
| 20-29 years old | 483 (13.35) | 481 (14.16) | 2 (0.90) |
| 30-39 years old | 943 (26.07) | 937 (27.59) | 6 (2.71) |
| 40-49 years old | 825 (22.81) | 810 (23.85) | 15 (6.79) |
| 50-59 years old | 597 (16.51) | 575 (16.93) | 22 (9.95) |
| 60-69 years old | 328 (9.07) | 284 (8.36) | 44 (19.91) |
| 70-79 years old | 181 (5.00) | 130 (3.83) | 51 (23.08) |
| 80-89 years old | 114 (3.15) | 50 (1.47) | 64 (28.96) |
| 90 years old or more | 26 (0.72) | 10 (0.29) | 16 (7.24) |
| Gender |  |  |  |
| Male | 1701 (47.03) | 1580 (46.53) | 121 (54.75) |
| Female | 1916 (52.97) | 1816 (53.47) | 100 (45.25) |
| Race/Color |  |  |  |
| Asian | 233 (6.44) | 204 (6.01) | 29 (13.12) |
| White | 1176 (32.51) | 1113 (32.77) | 63 (28.51) |
| Unknown | 699 (19.33) | 660 (19.43) | 39 (17.65) |
| Indigenous | 5 (0.14) | 5 (0.15) | 0 (0.00) |
| Brown | 1249 (34.53) | 1176 (34.63) | 73 (33.03) |
| Black | 255 (7.05) | 238 (7.01) | 17 (7.69) |
| Education<sup>a</sup> |  |  |  |
| Illiterate | 65 (1.80) | 47 (1.38) | 18 (8.14) |
| iES (1-4 grade) | 135 (3.73) | 117 (3.45) | 18 (8.14) |
| cES (4 grade) | 75 (2.07) | 69 (2.03) | 6 (2.71) |
| iES (5-8 grade) | 165 (4.56) | 150 (4.42) | 15 (6.79) |
| cES (8 grade) | 173 (4.78) | 160 (4.71) | 13 (5.88) |
| iHS | 146 (4.04) | 136 (4.00) | 10 (4.52) |
| cHS | 971 (26.85) | 947 (27.89) | 24 (10.86) |
| iHE | 88 (2.43) | 87 (2.56) | 1 (0.45) |
| cHE | 604 (16.70) | 599 (17.64) | 5 (2.26) |
| Unknown | 1195 (33.04) | 1084 (31.92) | 111 (50.23) |
| Hospitalization | 3617 (100.00) | 3396 (93.89) | 221 (6.11) |
| yes | 306 (8.46) | 210 (6.18) | 96 (43.44) |
| no | 2496 (69.01) | 2388 (70.32) | 108 (48.87) |
| unknown | 815 (22.53) | 798 (23.50) | 17 (7.69) |
| Travelling History | 3617 (100.00) | 3396 (93.89) | 221 (6.11) |
| Brazil |  |  |  |
| yes | 236 (6.52) | 231 (6.80) | 5 (2.26) |
| no | 2500 (69.12) | 2322 (68.37) | 178 (80.54) |
| unknown | 881 (24.36) | 843 (24.82) | 38 (17.19) |
| International |  |  |  |
| yes | 2 (0.06) | 2 (0.06) | 0 (0.00) |
| no | 2435 (67.32) | 2269 (66.81) | 166 (75.11) |
| unknown | 1180 (32.62) | 1125 (33.13) | 55 (24.89) |
| Symptom | 3617 (100.00) | 3396 (93.89) | 221 (6.11) |
| Fever |  |  |  |
| yes | 2077 (57.42) | 1928 (56.77) | 149 (67.42) |
| no | 1478 (40.86) | 1406 (41.40) | 72 (32.58) |
| unknown | 62 (1.71) | 62 (1.83) | 0 (0.00) |
| Respiratory Distress |  |  |  |
| yes | 956 (26.43) | 814 (23.97) | 142 (64.25) |
| no | 2601 (71.91) | 2522 (74.26) | 79 (35.75) |
| unknown | 60 (1.66) | 60 (1.77) | 0 (0.00) |
| Cough |  |  |  |
| yes | 2237 (61.85) | 2096 (61.72) | 141 (63.80) |
| no | 1317 (36.41) | 1238 (36.45) | 79 (35.75) |
| unknown | 63 (1.74) | 62 (1.83) | 1 (0.45) |
| Runny nose |  |  |  |
| yes | 1249 (34.53) | 1212 (35.69) | 37 (16.74) |
| no | 2305 (63.73) | 2122 (62.49) | 183 (82.81) |
| unknown | 63 (1.74) | 62 (1.83) | 1 (0.45) |
| Sore Throat |  |  |  |
| yes | 975 (26.96) | 955 (28.12) | 20 (9.05) |
| no | 2578 (71.27) | 2378 (70.02) | 200 (90.50) |
| unknown | 64 (1.77) | 63 (1.86) | 1 (0.45) |
| Diarrhea |  |  |  |
| yes | 465 (12.86) | 450 (13.25) | 15 (6.79) |
| no | 3087 (85.35) | 2882 (84.86) | 205 (92.76) |
| unknown | 65 (1.80) | 64 (1.88) | 1 (0.45) |
| Headache |  |  |  |
| yes | 1628 (45.01) | 1588 (46.76) | 40 (18.10) |
| no | 1925 (53.22) | 1745 (51.38) | 180 (81.45) |
| unknown | 64 (1.77) | 63 (1.86) | 1 (0.45) |
| Comorbidity | 3617 (100.00) | 3396 (93.89) | 221 (6.11) |
| Pulmonary |  |  |  |
| yes | 168 (4.64) | 145 (4.27) | 23 (10.41) |
| no | 3385 (93.59) | 3189 (93.90) | 196 (88.69) |
| unknown | 64 (1.77) | 62 (1.83) | 2 (0.90) |
| Cardiac |  |  |  |
| yes | 726 (20.07) | 602 (17.73) | 124 (56.11) |
| no | 2830 (78.24) | 2734 (80.51) | 96 (43.44) |
| unknown | 61 (1.69) | 60 (1.77) | 1 (0.45) |
| Kidney |  |  |  |
| yes | 43 (1.19) | 28 (0.82) | 15 (6.79) |
| no | 3511 (97.07) | 3306 (97.35) | 205 (92.76) |
| unknown | 63 (1.74) | 62 (1.83) | 1 (0.45) |
| Diabetes |  |  |  |
| yes | 345 (9.54) | 276 (8.13) | 69 (31.22) |
| no | 3209 (88.72) | 3058 (90.05) | 151 (68.33) |
| unknown | 63 (1.74) | 62 (1.83) | 1 (0.45) |
| Smoking |  |  |  |
| yes | 60 (1.66) | 50 (1.47) | 10 (4.52) |
| no | 3491 (96.52) | 3281 (96.61) | 210 (95.02) |
| unknown | 66 (1.82) | 65 (1.91) | 1 (0.45) |
| Obesity |  |  |  |
| yes | 156 (4.31) | 141 (4.15) | 15 (6.79) |
| no | 3378 (93.39) | 3174 (93.46) | 204 (92.31) |
| unknown | 83 (2.29) | 81 (2.39) | 2 (0.90) |

<sup>a</sup>iES= incomplete Elementary school; cES= complete Elementary school; iHS= incomplete High school; cHS= complete High School; iHE= incomplete Higher Education; cHE= complete Higher Education

Some of the variables have unknown values due to lack of information. Instances with such a characteristic were kept in the dataset as an *unknown* category, as there was no decrease in the performance of the models due to their presence.

According to recent studies related to COVID-19, older age and the presence of comorbidities are aggravating factors that can contribute to the disease severity. In addition, the presence of two or more clinical symptoms was considered important in the COVID-19 severity [18]. Thus, in order to add more knowledge to the dataset, additional variables were developed, namely: (i) sum of the comorbidities presented by the patient, (ii) sum of the symptoms presented by the patient and (iii) indicative if the patient has more than 60 years old. These new variables provide information that can contribute to predict the outcome of a new COVID-19 patient. They are calculated based on already existing variables from Tables 1 and 2. Our final datasets contains 24 independent variables and the target variable, represented by the disease outcome^1^: cure or death.

Tables 1 and 2 also present the distribution of the two classes, i.e., cure and death. It can be seen that we have imbalanced data, as the number of deaths corresponds only to 8.21% and 6.11% of the samples in the training and validation datasets, respectively. This difference can be a problem for machine learning models, making it difficult to predict samples of the minority class. Strategies to deal with this situation are often used, such as weighting and resampling [19]. We employed an oversampling strategy, increasing the number of death samples in order to obtain a balanced dataset. A simple procedure based on randomly picking samples with replacement was performed.

Figure 1 shows the correlation heatmap for the training dataset variables. It can be observed in the last line that some of the variables have a high correlation with the target variable, i.e., the disease outcome. They include age, respiratory distress, sum of commorbidities, hospitalization and age greater equal 60 years old. Similar correlations were found by [20] regarding age and chronic diseases.

**Figure 1:**
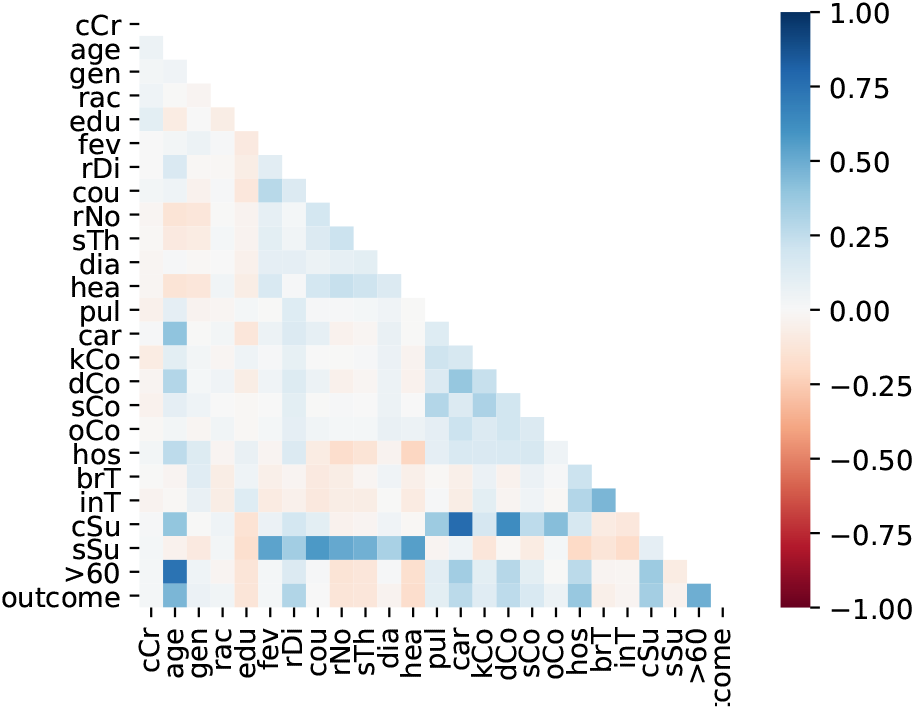
Variables correlation

### 2.2 Machine Learning Models

Machine Learning [21] is a research area which is concerned with the question of how to construct computer programs that automatically improve with experience. Recently, many successful machine learning applications have been developed.

Machine learning algorithms have proven to be of great practical value in a variety of application domains such as medical domain. They are especially useful in problems where databases may contain valuable implicit regularities that can be discovered automatically, e.g., to analyze outcomes of medical treatments from patient databases.

A classification problem consists of identifying to which of a set of categories a new instance belongs, given a historical data used for training, which contains instances whose category membership is known. This type of problem is solved through supervised learning.

In this paper, some supervised machine learning algorithms have been applied to a dataset having information from patients who tested positive for COVID-19 aiming to create computational models able to predict their disease outcome.

#### 2.2.1 1Logistic Regression (LR)

Logistic Regression (also called Logit Regression) is commonly used to estimate the probability that an instance belongs to a certain class. If the estimated probability is greater than 50%, then the model predicts that the instance belongs to that class (positive class), or else it does not (negative class). This turns LR model into a binary classifier, however it can be generalized to support multiple classes [22]. A LR model calculates a weighted sum of the input features (plus a polarization term), but instead of generating the result directly, a sigmoid (or logistic) function is applied. The sigmoid function (S-format) shows a number between 0 and 1. Once the LR model has estimated the probability that instance x belongs to the positive class, it can easily make its prediction.

#### 2.2.2 Linear Discriminant Analysis (LDA)

Linear Discriminant Analysis is a technique for calculating a linear transformation. It takes into account class information for calculating the transformation so that the separation between classes is maximum in the new coordinate space. LDA is also known as Fisher’s Linear Discriminant [23], due to the work of R. Fisher. The transformation of LDA is calculated so that the new coordinate system produces data with maximum variance between classes and minimum intraclass variance. LDA can be very effective in identifying large linearly separable problems.

#### 2.2.3 Naive Bayes (NB)

The Naive Bayes is an example of an induced classifier based on strong and unrealistic assumption: all the variables are considered to be conditionally independent given the value of the class variable. Consequently, a NB classifier is automatically achieved by only inducing the numerical parameters of the model. To this end, only information about the variables and their corresponding values are needed to estimate probabilities, leading to a computational time complexity that is linear with respect to the amount of training instances. NB is also space efficient, requiring only the information provided by two-dimensional tables, in which each entry corresponds to a probability estimated for a given value of a particular variable. According to [24], NB has provided good results on several domains.

#### 2.2.4 K-Nearest Neighbors (KNN)

The K-Nearest Neighbors algorithm is based on the concept of neighborhood, in which the neighbors are similar. Thus, it is possible to classify the elements of an n-dimensional space into *K* sets. This parameter *K* represents the number of neighbors and it is defined by the user in order to obtain a better classification. Classification is calculated based on a vote of the *K*-neighbors closest to each point (each instance of data or training example is viewed as points in space). According to [25], the classifier can get good results when there is lot of data in a low dimension (domains with few variables). However, in large dimensional spaces, usually the closest neighbors are distant.

#### 2.2.5 Decision Trees (DT)

Classification and Regression Tree (CART) is an algorithm to train Decision Trees (DT) [22]. A decision tree returns a response after executing a test sequence and it is considered one of the most successful methods of machine learning [25]. The CART algorithm works by first splitting the training set into two subsets using a single feature *k* and a threshold *t*_*k*_. It searches for the pair (*k, t*_*k*_) that produces the purest subsets (weighted by their size). Once the CART algorithm has successfully split the training set in two, it splits the subsets using the same logic, then the sub-subsets, and so on, recursively. It stops recursing once it reaches the maximum depth or if it cannot find a split that will reduce impurity.

#### 2.2.6 XGBOOST (XGB)

XGBoost (eXtreme Gradient Boosting) is an implementation of stochastic gradient boosting. This implementation is computationally efficient with many options and is available as a package for the main data science software languages [26]. The XGB library implements the gradient boosting decision tree algorithm. It was designed to be highly efficient, flexible and portable. Gradient boosting is an approach where new models are created that predict the residuals or errors of prior models and then added together to make the final prediction. It is called gradient boosting because it uses a gradient descent algorithm to minimize the loss when adding new models. XGB provides a parallel tree boosting that solves many data science problems in a fast and accurate way. This approach supports both regression and classification predictive modeling problems.

#### 2.2.7 Support Vector Machine (SVM)

Support Vector Machine (SVM) is a supervised learning method which became popular in some years ago for solving problems in classification, regression, and novelty detection [27]. The SVM approaches the problem of finding one solution that separate the classes exactly from the training data set through the concept of the margin, which is defined to be the smallest distance between the decision boundary and any of the samples. SVM constructs a decision boundary (maximum margin separator) with the greatest possible distance to example points. The idea of SVM is to focus on points more important than others that lead to the best generalization. For this, a linear separation in hyperplane is created, even if the data are not separable linearly in the original input space, because they can incorporate the data in a space of superior dimension, using kernel trick. The linear dimension separator is actually nonlinear in the original space.

### 2.3 Evaluation Metrics

In this study, we evaluate the performance of each of the learning models in terms of accuracy, Receiver Operating Characteristic curve and area under the curve, precision, recall, Precision-Recall curve and area under the curve, F1-score and finally the confusion matrix. These metrics are detailed in the following.

1. Confusion Matrix: in a binary classification, the result on a test set is often displayed as a two-dimensional *confusion matrix* with a row and column for each class. Each matrix element shows the number of test examples for which the actual class is the row and the predicted class is the column. Good results correspond to large numbers down the main diagonal and small, ideally zero, off-diagonal elements [28]. The scheme of a confusion matrix is illustrated below.

2. Accuracy: it is the ratio of the number of correct predictions to the total number of samples. It works well when there are equal number of samples belonging to each class. However, accuracy is misleading for skewed class distribution since correct predictions for the minority class can fully ignored. It can be given by:

|  |  | predicted class |  |
| --- | --- | --- | --- |
|  |  | cure | death |
| actual class | cure | TN | FP |
|  | death | FN | TP |

TN=true negative, FP=false positive
FN=false negative, TP=true positive

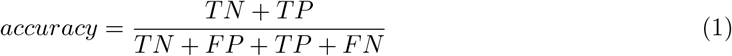
3. Precision: also known as the positive predictive value, precision is defined as the proportion of positive examples that are truly positive. A precise model will only predict the positive class in cases very likely to be positive. This metric can be calculated by following formula:

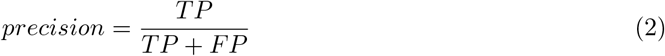
4. Recall: also known as sensitivity, is a measure of how complete the results are. A model with high recall captures a large portion of the positives examples, meaning that it has wide breadth. It is calculated as:

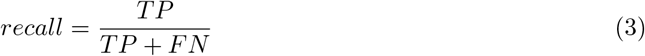
5. F1-score: this metric seeks a balance between precision and recall and represents an interesting metric when there is an uneven class distribution. It is given by the harmonic mean of precision and recall:

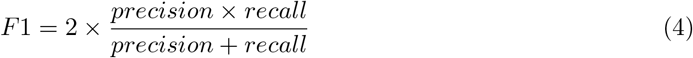
6. Receiver Operating Characteristic (ROC) curve: ROC curves are a graphical technique for evaluating the performance of a binary classifier at different classification thresholds. These curves depict the performance of a classifier without regard to class distribution or error costs. They plot the TP rate on the vertical axis against the FP rate on the horizontal axis.
7. Precision-Recall (PR) curve: a precision-recall curve shows the relationship between precision (positive predictive value) and recall (sensitivity) for every possible cut-off. It represents an alternative to a ROC curve and shows a good choice to compare models when facing imbalanced datasets.The main difference between ROC curves and PR curves is that the number of TN results is not used to make a PR curve.
8. Area Under the Curve (AUC): AUC measures the entire two dimensional area underneath an entire curve. Therefore, it gives an aggregate measure of performance in a single value. AUC ranges from 0.0 to 1.0; a model with predictions 100% correct has an AUC of 1.0 while one whose predictions are 100% wrong gives an AUC of 0.0. We use AUC values for ROC and PR curves in our experiments.

## 3 Results

Our experimental design involves two main parts. The first, named *Experiment 1*, consists of a series of repeated tests using the training dataset, while the second, *Experiment 2*, performs a final test by using both the training and validation datasets.

The first experiment includes 4, 826 patients (46.25% male and 53.75% female), distributed in different age groups, educational level, race/color, hospitalization and travelling history. Of the total number of patients, 91.79% cured and 8.21% deceased. Table 1 shows 7 symptoms and 6 comorbidities present/absent among those who cured or died. The second experiment includes a new dataset containing 3, 617 patients (47.03% male and 52.97% female) of which 93.89% cured and 6.11% died (Table 2).

Both datasets present similar distributions among the different variables’ categories. However, it is possible to observe the increase in the number of confirmed (closed) cases, since the validation dataset comprises only 7 days and has only 25% fewer samples than the training dataset that corresponds to a period of 75 days. These numbers reflect the rapid progress of cases in Brazil.

### 3.1 Experiment 1

Experiment 1 is developed to evaluate the performance of the different prediction models under a series of repeated tests using different partitions of the training dataset (Table 1). The idea underlying this experiment is illustrated in Figure 2. A 70-30 split is performed in the dataset through a random, but stratified procedure. The 70-part is used for training by a 10-fold cross validation, with oversampling applied only in the training folds (9 of them), generating the training results, i.e., an estimate of the performance of the models. Once we have the trained models, the 30-part is used for validation, leading to the test results. This procedure is repeated 33 times and the results are reported in Figures 3 and 4. Grid search was used in order to find the best hyper-parameters for the models.

**Figure 2:**
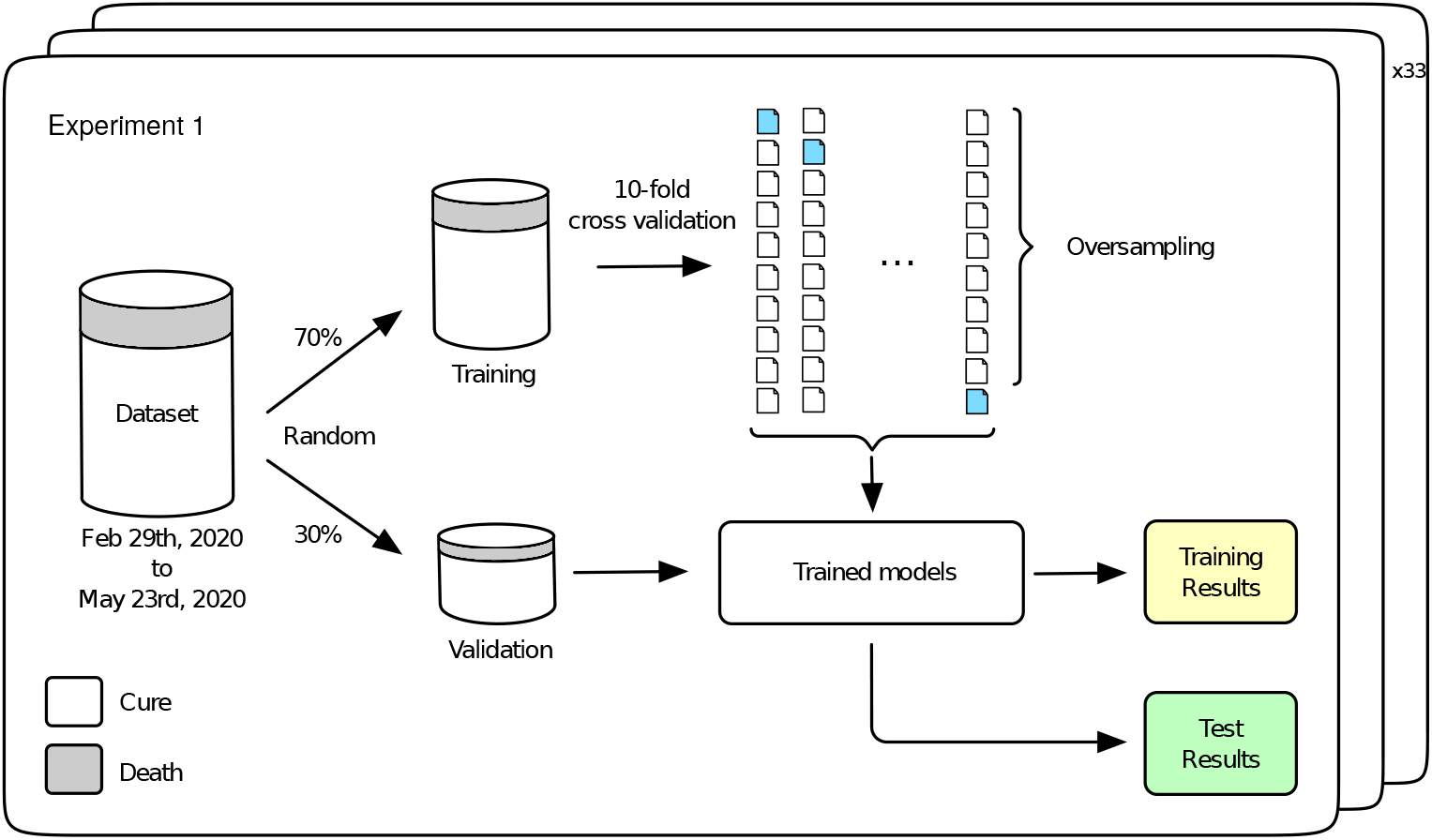
Experiment 1

**Figure 3:**
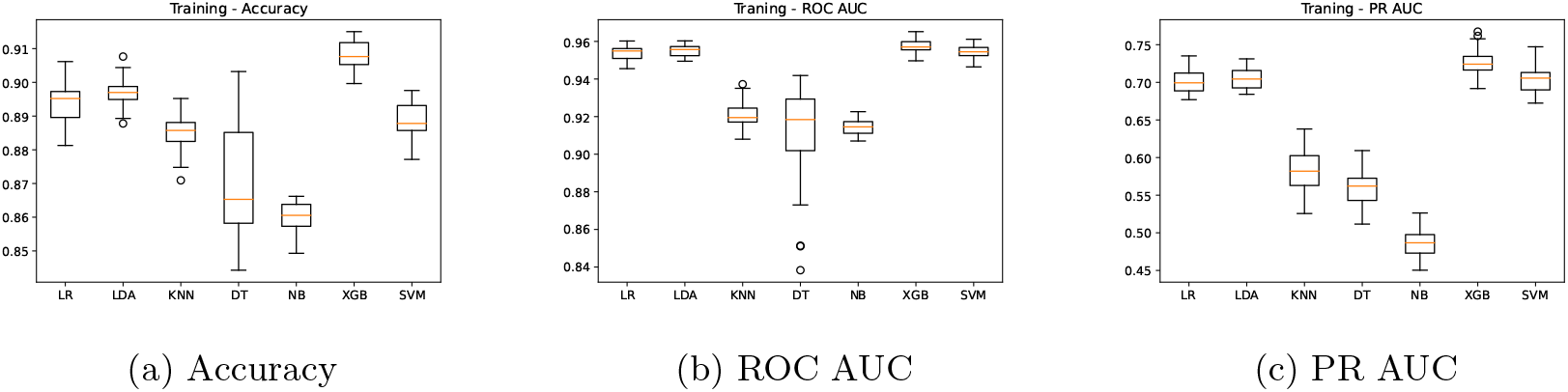
Training Results - Algorithms Comparison

**Figure 4:**
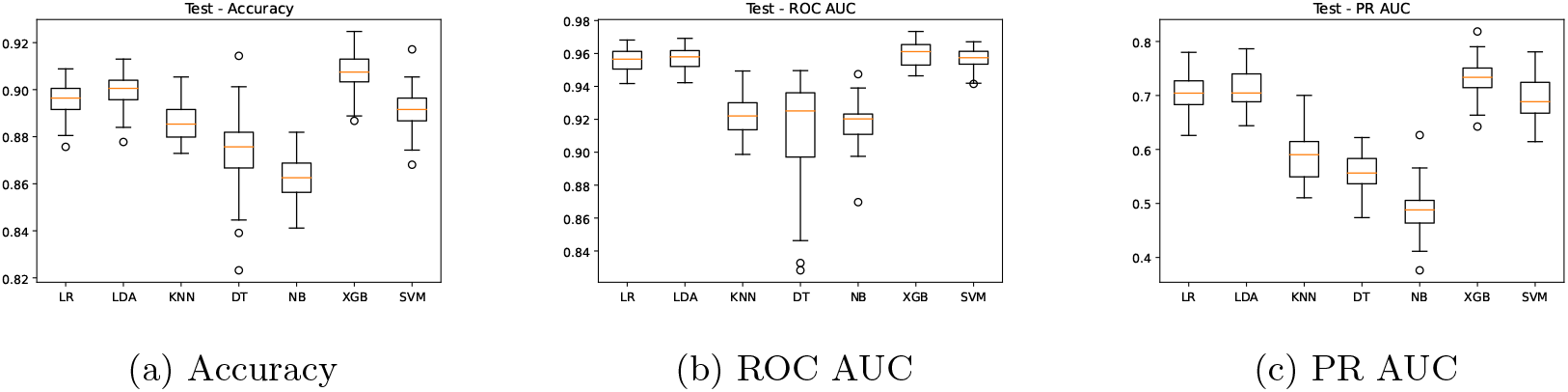
Test Results - Algorithms Comparison

Figure 3 shows summarized training results for the seven prediction models regarding accuracy, ROC AUC and PR AUC metrics. A higher performance for the three metrics is achieved by LR, LDA, XBG and SVM models, with ROC AUC mean values of 0.95, 0.95, 0.96 and 0.95, respectively (Figure 3b). On the other hand, models KNN, DT and NB present a ROC AUC of 0.92, 0.91 and 0.91, respectively. In Figure 3c, we present the precision-recall AUC, which does not consider the true negative results, giving a higher importance to the minority class. The PR AUC mean value is around 0.7 for the best models (LR=0.70, LDA=0.70, XGB=0.72 and SVM=0.70), while models KNN, DT and NB have 0.58, 0.55 and 0.48 mean values. As mentioned before, in this study the accuracy is not the best metric to compare the different models. According to Figure 3a, LR, LDA, XBG and SVM models achieve accuracy mean values of 0.89, 0.90, 0.91, 0.89, while for KNN, DT and NB we have 0.88, 0.87 and 0.86, respectively. As we can see, although the accuracy values do not present significant differences among the models, the other two metrics (ROC AUC and PR AUC) make performance differences more evident. The DT model has a lower robustness due to a higher dispersion while the NB model presents the worse performance.

In Figure 4, we show the summarized test results using the 30-part from the split and the trained models. This evaluation aims to check whether the models were able to learn from the data and achieve good predictions with unseen data. A similar behavior is observed for all metrics, keeping LR, LDA, XGB and SVM as the best models. The accuracy remains high, with mean values above 0.89 for these models (Figure 4a), while the ROC AUC is around 0.95 (Figure 4b). A PR AUC of 0.7 (4c) on average is also achieved by the test results. These results verify the learning capacity of the models and the absence of overfitting, as there was no significant drop in performance in the test results.

### 3.2 Experiment 2

The second part of our experimental design concerns a validation test using new data from Table 2. For that, algorithms were trained using all data from the training dataset, i.e., closed cases included in the database until May 23rd, 2020 while prediction was performed for the validation dataset. The main difference of experiments 1 and 2 rely on the validation data being sequential samples for *Experiment 2*, while in *Experiment 1* validation sets correspond to samples randomly selected from the whole dataset. This temporal aspect may show importance since future predictions will be made for new patients (sequential samples) being tested positive for COVID-19.

The general conception of *Experiment 2* is shown in Figure 5. A training step is performed using the whole training dataset through 10-fold cross validation and oversampling, producing the training results. With the trained models in hands, a validation step is developed, making predictions for the whole validation dataset and leading to the final test results. Training results are reported on Table 3 while final tests results are shown through Figures 6 and 7. Again, grid search was used in order to find the best hyper-parameters for the models.

**Table 3:**
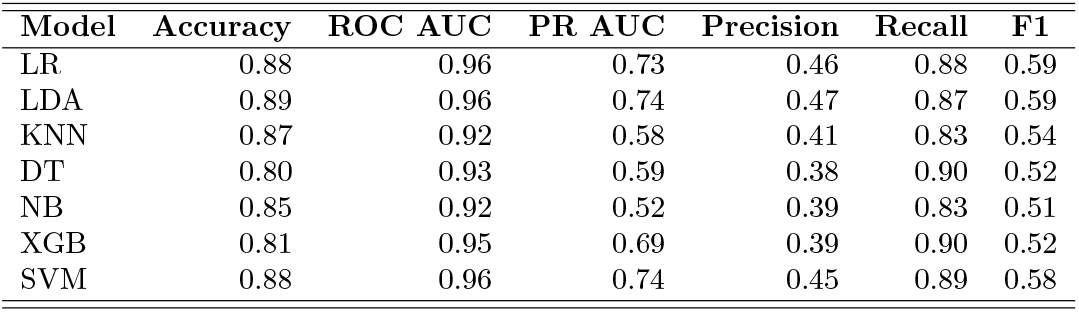
Experiment 2 Training Models Performance

**Figure 5:**
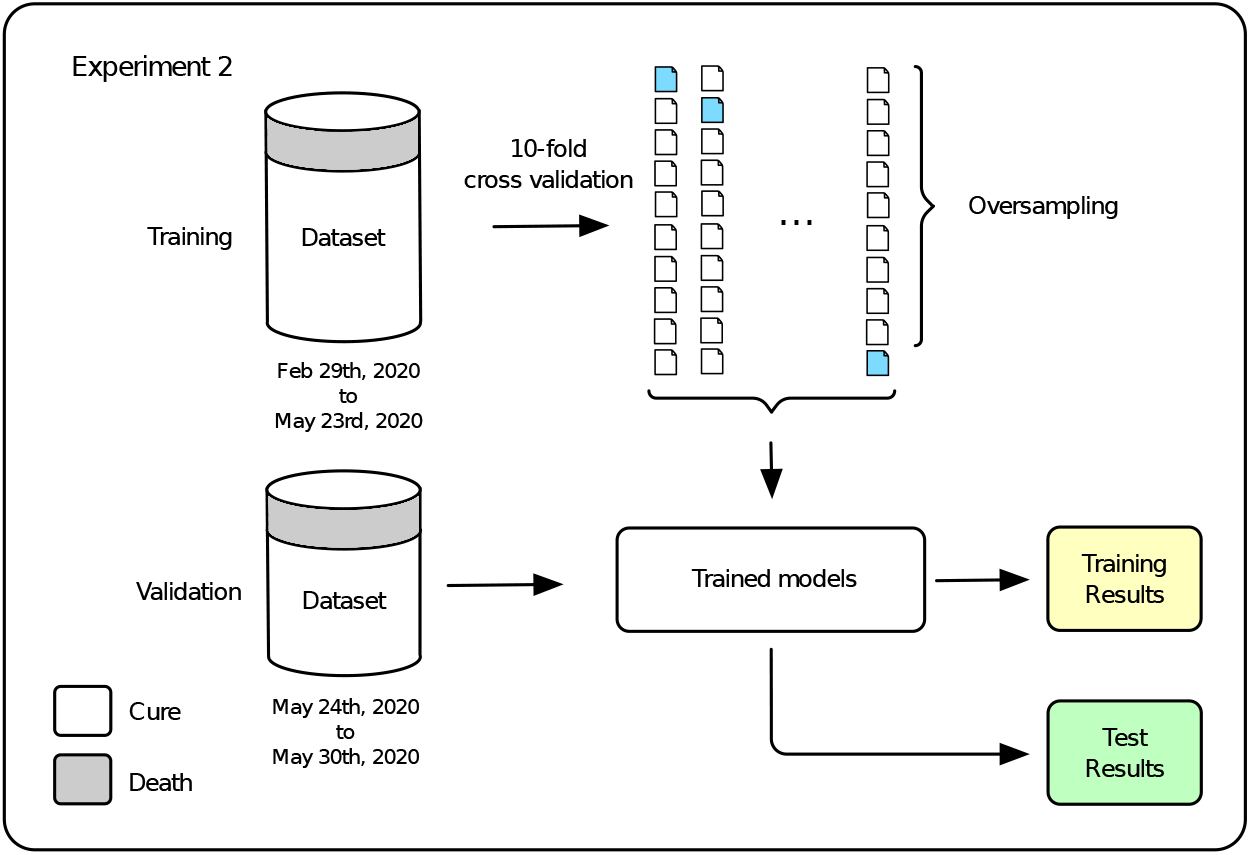
Experiment 2

**Figure 6:**
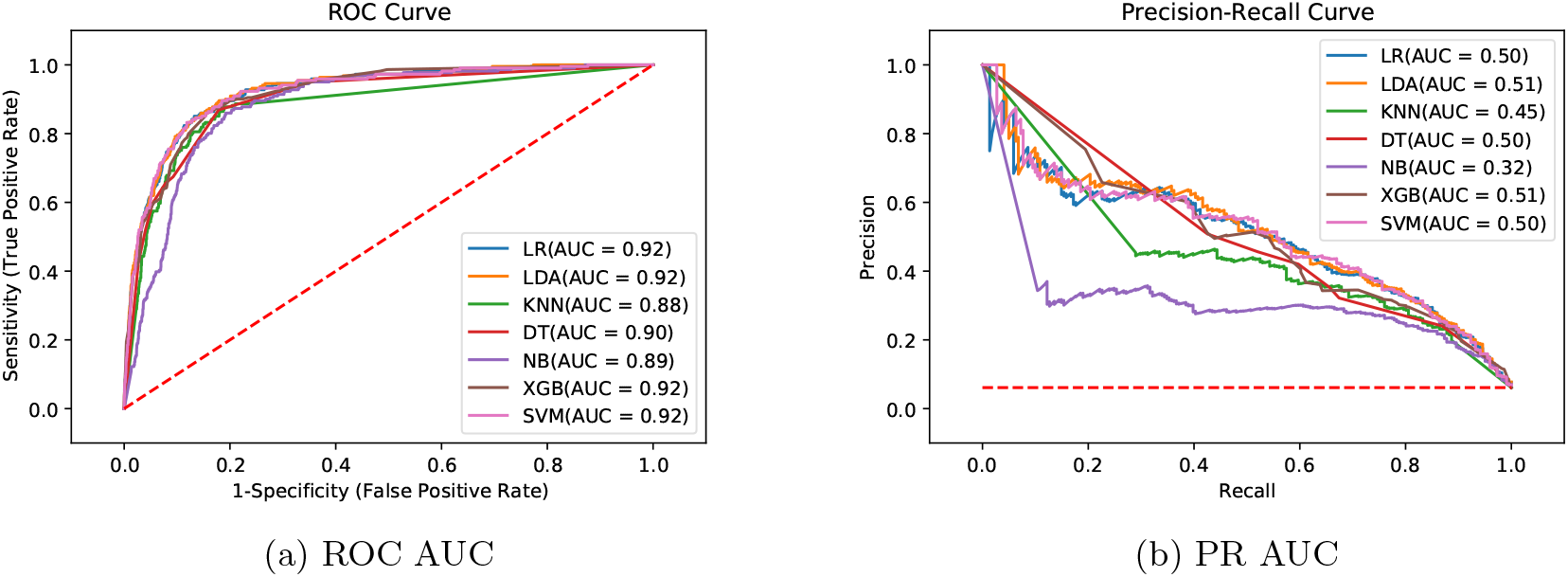
Test Results - Algorithms Comparison

**Figure 7:**
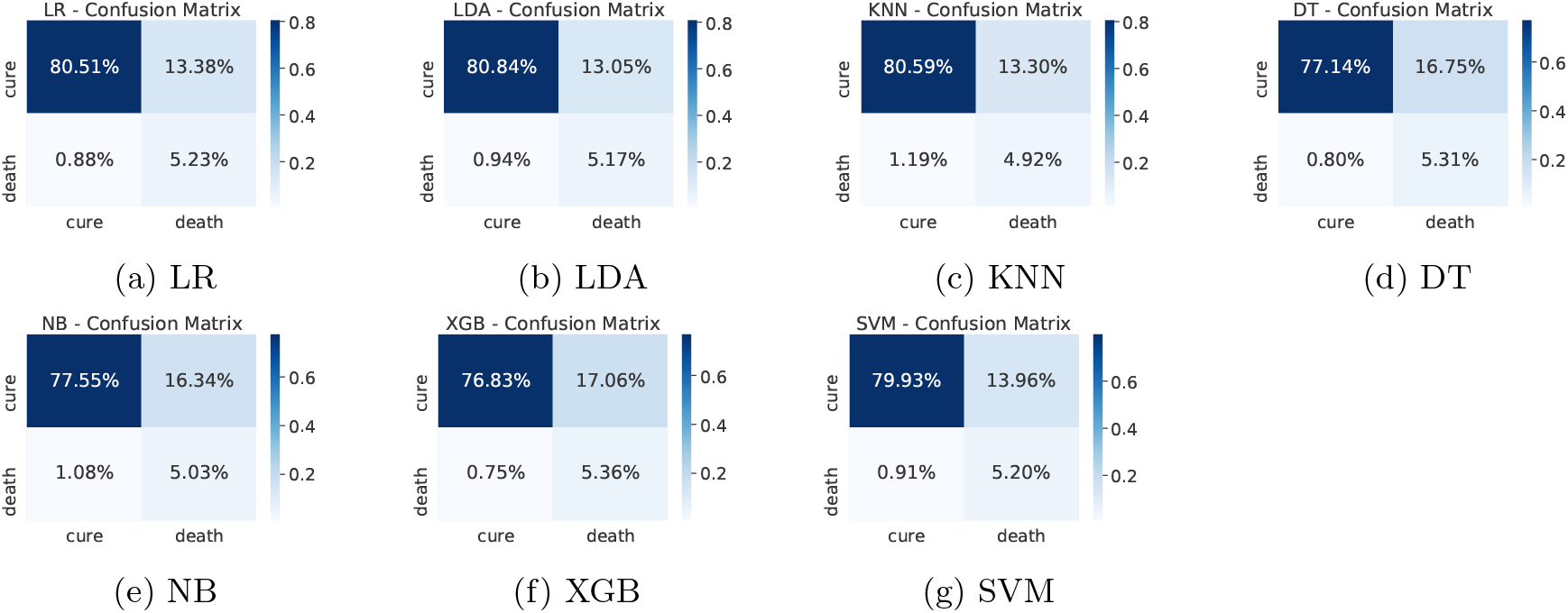
Test Results - Confusion Matrices

The training results of *Experiment 2* are detailed in Table 3, presenting the following metrics: accuracy, ROC AUC, PR AUC, Precision, Recall and F1-score. All metric results are consistent with the training results of the former experiment. Again, the best models regarding ROC AUC and PR AUC are LR, LDA, XBG and SVM. It is important to mention that the minority class has high importance in our application, leaving the accuracy as a secondary metric. One can note that although KNN and NB models have high accuracy values, they present the worst recall values.

On the analysis of the test results, it can be seen in Figure 6 the representation of the ROC curve (Figure 6a) and PR curve (Figure 6b). Detailed results for the whole set of metrics are shown in Table 4. Table 5 presents the best parameters used in these final experiments. Comparing the ROC AUC values from Table 3 and Figure 6a, a slight drop in values can be noted for all models, with a decrease of 0.037 on average. This behaviour is expected since we are using completely new data, but no significant difference that indicates overfitting or a poor learning step. Most of the models have a very close performance, making it difficult to select one as the best model. From Figure 6b, a greater difference among the models can be noted for the PR AUC metric. Models KNN and NB show a clearly underperformance compared to the other models. In general, this metric shows an inferior performance in the tests in relation to the training step for all models. This can be due to the difference on the number of samples in the minority class. While the training dataset has 8.21% of the samples in death class, the validation dataset is even more imbalanced, with only 6.11% of the samples belonging to that class.

**Table 4:** Experiment 2 Test Models Performance

| Model | Accuracy | ROC AUC | PR AUC | Precision | Recall | F1 |
| --- | --- | --- | --- | --- | --- | --- |
| LR | 0.86 | 0.92 | 0.50 | 0.28 | 0.86 | 0.42 |
| LDA | 0.86 | 0.92 | 0.51 | 0.28 | 0.85 | 0.43 |
| KNN | 0.85 | 0.88 | 0.45 | 0.27 | 0.80 | 0.40 |
| DT | 0.82 | 0.90 | 0.50 | 0.24 | 0.87 | 0.38 |
| NB | 0.82 | 0.89 | 0.32 | 0.23 | 0.82 | 0.37 |
| XGB | 0.82 | 0.92 | 0.51 | 0.24 | 0.88 | 0.38 |
| SVM | 0.85 | 0.92 | 0.50 | 0.27 | 0.85 | 0.41 |

**Table 5:** Hyperparameter Tuning in Validation Tests

| Model | Parameters |
| --- | --- |
| LR | C = 100, penalty = l2, solver = liblinear, multi_class = ovr |
| LDA | solver = svd |
| KNN | n_neighbors = 19, weights = distance, metric = euclidean |
| DT | max_depth = 3 |
| NB | - |
| XGB | max_depth = 3, n_estimators = 100, learning_rate = 0.01, subsample = 1 |
| SVM | kernel = rbf, C = 10, gamma = 0.0001 |

Moreover, it is important to mention that unlike ROC AUC, whose baseline is 0.5 (random guess classifier), in PR AUC the baseline is dependent on the application itself. In the case of this work, a random estimator would have a PR AUC of 0.06 (6.11% in death class, see the horizontal line in Figure 6b) and therefore, values around 0.5 are definitely a substantial increase.

From Table 4 and Figure 6b, we can observe that although the results show a high recall, they present a relatively low precision. This means the minority class (death) is well detected but the model also include points of the other class (cure) in it. This fact is confirmed by the confusion matrices introduced in Figure 7. We can note an expressive amount of false positives samples, represented in the upper right corner. False positives concern patients who cured but were wrongly classified as deaths by the models. It is possible to note that all models present a similar behavior regarding this wrong prediction. Aiming to find an explanation for this behavior, we decided to analyse the characteristics of these samples looking for similarities.

Analysis of the characteristics of false positive patients shows that such patients had at least one of the following critical situations: *>* 60 years old, respiratory distress, some comorbidity and hospitalization (see Table 8). It is tempting to speculate that, in a way, this percentage of patients predicted in the model could be those with the most critical condition, but that due to early and effective care were cured. This could even be a positive aspect of the models prediction, since it is important to identify severe cases which deserve special care. Unfortunately, it is not possible to confirm such a hypothesis since the database does not provide information to differentiate mild and severe cases from those who have been cured. In an attempt to show that such patients may have been critical cases, Table 8 shows the percentage of patients who have the characteristics most related to the chance of death according to the calculation of odds ratio from Tables 6 and 7.

**Table 6:** Odds ratio for training dataset

| Condition | OR | 95% CI | P |
| --- | --- | --- | --- |
| >60 years old | 25.30 | (19.83-32.34) | <0.0001 |
| Fever | 1.16 | (0.93-1.14) | = 0.1835 |
| Respiratory Distress | 8.34 | (6.64-10.48) | <0.0001 |
| Cough | 1.00 | (0.81-1.25) | = 0.9710 |
| Runny nose | 0.27 | (0.21-0.35) | <0.0001 |
| Sore Throat | 0.26 | 0.18-0.36() | <0.0001 |
| Diarrhea | 0.61 | (0.41-0.88) | = 0.0090 |
| Headache | 0.21 | (0.16-0.27) | <0.0001 |
| Pulmonary Disease | 3.53 | (2.51-4.96) | <0.0001 |
| Cardiac Disease | 6.48 | (5.22-8.05) | <0.0001 |
| Kidney | 12.96 | (7.10-23.60) | <0.0001 |
| Diabetes | 7.70 | (6.01-9.85) | <0.0001 |
| Smoking | 10.08 | (6.40-15.80) | <0.0001 |
| Obesity | 2.14 | (1.49-3.07) | <0.0001 |
| Hospitalization | 23.88 | (18.50-30.82) | <0.0001 |

**Table 7:**
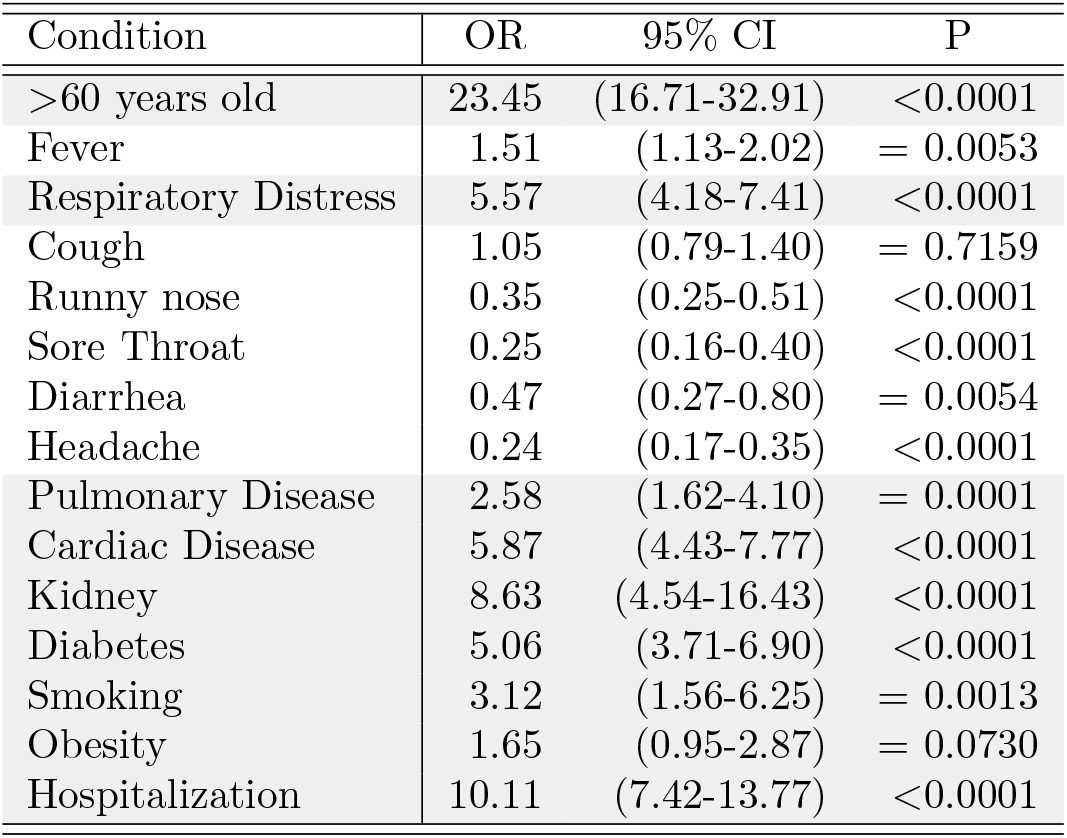
Odds ratio for validation dataset

| Condition | OR | 95% CI | P |
| --- | --- | --- | --- |
| >60 years old | 23.45 | (16.71-32.91) | <0.0001 |
| Fever | 1.51 | (1.13-2.02) | = 0.0053 |
| Respiratory Distress | 5.57 | (4.18-7.41) | <0.0001 |
| Cough | 1.05 | (0.79-1.40) | = 0.7159 |
| Runny nose | 0.35 | (0.25-0.51) | <0.0001 |
| Sore Throat | 0.25 | (0.16-0.40) | <0.0001 |
| Diarrhea | 0.47 | (0.27-0.80) | = 0.0054 |
| Headache | 0.24 | (0.17-0.35) | <0.0001 |
| Pulmonary Disease | 2.58 | (1.62-4.10) | = 0.0001 |
| Cardiac Disease | 5.87 | (4.43-7.77) | <0.0001 |
| Kidney | 8.63 | (4.54-16.43) | <0.0001 |
| Diabetes | 5.06 | (3.71-6.90) | <0.0001 |
| Smoking | 3.12 | (1.56-6.25) | = 0.0013 |
| Obesity | 1.65 | (0.95-2.87) | = 0.0730 |
| Hospitalization | 10.11 | (7.42-13.77) | <0.0001 |

**Table 8:** Critical conditions present in false positive patients

| Condition | LR | LDA | KNN | DT | NB | XGB | SVM | AVG |
| --- | --- | --- | --- | --- | --- | --- | --- | --- |
| >60 years old | 64.05 | 74.36 | 61.95 | 78.22 | 55.84 | 76.82 | 70.30 | 68.79 |
| Respiratory Distress | 59.30 | 51.69 | 54.68 | 36.30 | 35.70 | 37.44 | 51.49 | 46.66 |
| Comorbidity | 60.74 | 61.23 | 62.16 | 52.48 | 75.30 | 53.32 | 59.41 | 60.66 |
| Hospitalization | 34.30 | 33.05 | 29.94 | 34.65 | 20.64 | 34.04 | 34.46 | 31.58 |
| **At least one condition** | 100.00 | 100.00 | 98.34 | 100.00 | 90.52 | 100.00 | 100.00 | 98.41 |

Table 6 shows that the death chance is greater among COVID-19 patients over 60 years old. Respiratory distress and comorbidities such as kidney disease, diabetes, cardiac disease and obesity, as well as smoking, increase the likelihood of death from COVID-19. On the other hand, runny nose, sore throat, diarrhea and headache were less likely to occur in patients who deceased. Our results are similar to those reported by [29] in severe COVID-19 patients compared to non-severe patients, emphasizing a high probability of complication in patients with kidney disease. The validation cohort (Table 7) showed similar results, with the exception of the fever symptom, which was more likely to occur among patients who died compared to those who cured.

A brief review of other literature works is presented in the next section along with a discussion on how this work can help in the current scenario of COVID-19 in Brazil.

## 4 Discussion

Prediction models of the prognosis for a given disease have the main objective of supporting the physician’s decision-making about what is the best measure of patient referral, assisting in the screening of patients at high risk of progressing to severe disease.

Artificial intelligence models aiming to identify risk factors for prognostic prediction of severe COVID-19 have been developed using age, clinical characteristics, laboratory tests and chest imaging [30, 31, 32, 29, 33, 34].

A study using age, hypertension history and coronary heart disease showed good discriminatory power (AUC = 0.83) between COVID-19 surviving and non-surviving patients. The inclusion of biochemical data increased (AUC = 0.88) the discriminatory power [30]. Those results refer to the validation cohort consisting of 44 patients.

Blood parameters were also used to select predictive biomarkers of mortality through machine learning. Lactate dehydrogenase (LDH), lymphocytes and high-sensitivity C-reactive protein (hsCRP) proved to be good indicators for early predicting the degree of COVID-19 severity, with *>* 90% accuracy [32].

A machine learning study involving a cohort of 214 non-severe and 148 severe patients with COVID-19 found *>* 90% prediction accuracy for disease severity using symptom/comorbidities data.

The addition of biochemical data to symptoms/comorbidities achieved *>* 99% predictive accuracy. Therefore, it was suggested that symptoms and comorbidities can be used in an initial screening and the biochemical data inclusion could predict the severity degree and assist in the development of treatment plans [29].

Importantly, in relation to our study, the sample size in the aforementioned studies was limited, since they were carried out with data of the beginning of the pandemic. In fact, the sample size can influence the robustness of the models performance. Larger datasets provide a better training stage, potentially leading to better performance in prediction.

Although some studies have pointed out changes in blood parameters such as lymphopenia, neutrophilia and increased lactate dehydrogenase concentration [6, 35], as well as changes in the chest images [36] as good indicators of the disease severity, these data are not publicly available and were not included in our study due to lack of access. In future studies we intend to include such data and check if there is an improvement in models performance. In view of the costs and difficulties of performing laboratory and chest imaging exams for an alarmingly increasing number of patients, our study proves to be important in that it is able to differentiate those critically ill patients who need ICUs care using less complex approaches, that is, age, symptoms and comorbidities at the time of screening.

Brazil has an unified health system, namely Sistema Único de Saúde (SUS), that allows for almost universal health coverage across the country, despite regional inequalities. With the growing number of COVID-19 cases, the TeleSUS system [37] was implemented on May 2020 as a strategy to perform remote preclinical health care and avoid unnecessary travel and the exhaustion of on-site health services. In this context, our study could also assist in screening those who may need early care or hospitalization solely through reports of personal data, symptoms and comorbidities. This model can be applied in other localities that have overloaded healthcare systems. Moreover, this model can also help in understanding the upcoming demand for ICU beds, staff and other critical resources.

Finally, it is important to highlight that this study was based only on a database from a State (Espírito Santo) of Brazil, requiring application in other States, since regional variations can occur in a country with continental characteristics such as Brazil.

## Data Availability

The data used in this work is publicly available.

## Acknowledgement

We would also like to thank the Health Department of Espírito Santo for placing the COVID-19 database open access.

## Availability of data and material

The data used in this work is publicly available [17].

## Footnotes

1 The disease outcome is also referred to as a “class” throughout this text.

